# Patient Portal Activation Among Neurology Patients in Washington, DC

**DOI:** 10.64898/2026.04.08.26350061

**Authors:** Nicholas S. Streicher

**Affiliations:** Department of Neurology, MedStar Georgetown University Hospital, Georgetown University Medical Center, Washington, DC

## Abstract

**Background and Objectives:** Patient portals have become essential infrastructure for healthcare delivery following the 21st Century Cures Act, yet adoption remains inequitable. Understanding demographic and geographic determinants of portal activation is critical for addressing digital health disparities, particularly among neurology patients who face unique access barriers. We examined the demographic, geographic, and neighborhood-level factors associated with patient portal activation among neurology patients at multiple geographic scales in the Washington, DC metropolitan area.

**Methods:** We conducted a retrospective cohort study of 72,417 adult neurology patients seen at two academic medical centers sharing an electronic health record in Washington, DC (February 2021–February 2026). We examined portal activation using multivariable logistic regression and geographic analysis at four nested scales: the metropolitan catchment area, DC’s eight wards, individual census tracts (via geocoded patient addresses), and individual DC residents.

**Results:** Portal activation was 64.7% overall. Activation varied by race/ethnicity (Non-Hispanic White 76.1%, Non-Hispanic Black 57.0%, Non-Hispanic Asian 57.6%, Hispanic 55.0%) and geography (DC Ward 2: 82.0% vs. Ward 7: 48.0%). Ward-level educational attainment (r = 0.948), broadband access (r = 0.889), and income (r = 0.811) were strongly correlated with activation. Within individual wards, Non-Hispanic White patients activated at 84–91% while Non-Hispanic Black patients activated at 48–64%, demonstrating that neighborhood resources alone do not explain disparities.

**Discussion:** Patient portal activation is shaped by demographic, socioeconomic, and geographic factors operating at multiple levels. Persistent within-ward racial disparities indicate that geographically targeted interventions must be paired with culturally tailored approaches to achieve digital health equity.

## Introduction

Patient portals transitioned from convenience to necessity after the 21st Century Cures Act mandated real-time electronic access to test results in 2021.1 Portal access more than doubled in the past decade, with approximately three in five individuals accessing their records by 2022 and continued growth since.^2^ Activation rates vary widely across settings, reflecting differences in patient demographics, clinical context, and implementation strategy.^3,4,12^

For neurology patients managing chronic conditions, portal access enables critical functions: monitoring test results, adjusting medications, and maintaining communication with providers.^5^ Portal engagement has been associated with improved medication adherence and clinical outcomes.^14,15^ Neurological impairments create unique barriers—dementia affects comprehension and portal navigation, while conditions such as Parkinson’s disease and multiple sclerosis may impair the motor and visual function needed for digital access.^6^

While demographic predictors of portal non-activation are well characterized—age, race, income, education, and language have each been documented across multiple ^5,7,8,11,12,13^—what remains unknown is how these factors interact with neighborhood context at different geographic scales. Studies examining geographic access to neurologists have operated at the national level^23^ while portal disparities research has typically relied on single-center data or national surveys without sub-metropolitan resolution.^24^ The result is a body of evidence that identifies who is disadvantaged but not where the disadvantage concentrates or at what geographic scale it becomes most visible.

Washington, DC provides a strategic setting for examining digital health disparities. Two academic hospitals share an electronic health record system yet serve populations with vastly different demographic profiles. The District’s eight wards encompass dramatic socioeconomic variation, with median household income ranging from $ 53,000 to $ 162,000 and bachelor’s degree attainment from 23% to 88%.^20–22^ This variation creates a natural experiment for disentangling individual demographics, neighborhood context, and structural inequality as contributors to portal disparities.

Here, we examine portal activation among 72,417 neurology patients at four nested geographic scales: the metropolitan catchment area, DC’s eight wards, individual census tracts, and residential zip codes. This multi-level approach maps where the digital divide runs deepest and identifies which demographic and neighborhood factors concentrate non-activation.

## Methods

### Study Population and Setting

This retrospective cohort study identified 72,524 patients with at least one neurology encounter at two academic medical centers in the Washington, DC metropolitan area between February 2021 and February 2026; 107 (0.15%) were excluded for missing sex data, yielding an analytic cohort of 72,417. Hospital A is a university-affiliated academic medical center with affiliated outpatient sites and Hospital B is an academic tertiary care hospital; both share a common electronic health record. Patient demographics, portal activation status, and residential addresses were extracted from the electronic health record.

### Data Cleaning and Quality Assurance

Residential zip codes were standardized to 5-digit format; 420 ZIP+4 codes were truncated and 17 non-US postal codes were excluded from geographic analyses. Street addresses were parsed for quadrant designators (NW, NE, SE, SW) to improve ward assignment in multi-ward zip codes. Patients with P.O. Box addresses (n = 664) were retained in demographic analyses but excluded from geographic mapping. A small number of addresses listing homeless shelters (n = 18) were similarly handled. Race and ethnicity fields were harmonized from multiple coding schemes in the electronic health record into four consistent categories.

### Patient Portal Activation

The primary outcome was patient portal activation status (active vs. inactive) as recorded in the electronic health record at the time of data extraction. Overall activation was 64.7% (46,851 of 72,417 patients).

### Racial and Ethnic Classification

Race and ethnicity were classified into five mutually exclusive categories: Non-Hispanic White (n = 28,154, 38.9%), Non-Hispanic Black (n = 22,900, 31.6%), Hispanic (n = 3,600, 5.0%), Non-Hispanic Asian (n = 1,925, 2.7%), and Other/Unknown (n = 15,838, 21.9%). Patients with any Hispanic or Spanish ethnicity designation were classified as Hispanic regardless of race.

### Geographic Assignment

Patients resided across three jurisdictions: Maryland (n = 28,318, 39.1%), DC (n = 22,752, 31.4%), and Virginia (n = 18,441, 25.5%), with the remainder in other states. For DC residents, patients were assigned to one of eight wards based on their residential zip code and street address quadrant (NW, NE, SE, SW). For addresses in multi-ward zip codes lacking a quadrant designator, street name matching was used as a secondary assignment method. This combined approach assigned 22,536 of 22,752 DC patients (99.1%) to a specific ward; the 216 unassigned were predominantly P.O. Box addresses.

For the broader catchment area analysis, patient activation rates were aggregated by 5-digit zip code. Zip codes with fewer than 10 patients were excluded from visualization; those with fewer than 30 from correlation analyses. This yielded 130 zip codes representing 51,506 patients. Additionally, patient street addresses were geocoded to census tracts using the U.S. Census Bureau batch geocoder (Public_AR_Current benchmark). Of 71,724 eligible addresses (excluding P.O. boxes, homeless shelter addresses, and non-U.S. postal codes), 67,327 (93.9%) were successfully matched to a census tract, including 21,592 of 22,752 DC residents (94.9%) across all 206 DC census tracts. Geocoded and non-geocoded patients were demographically similar, suggesting minimal selection bias from geocoding failure.

### Neighborhood Socioeconomic Variables

Neighborhood-level characteristics were obtained from the American Community Survey (ACS) 2019–2023 5-year estimates via the tidycensus R package at the census tract level for DC, Maryland, and Virginia (3,879 tracts). Three variables were selected: median household income, educational attainment (percentage with bachelor’s degree or higher), and broadband internet subscription rate. For ward-level analyses, ACS estimates were obtained directly at the ward level, as DC wards are classified as state legislative districts (upper chamber) in Census geography. For individual-level models, ACS variables were linked directly to each patient’s geocoded census tract.

### Statistical Analysis

Multivariable logistic regression estimated the association between patient characteristics and portal activation across all three cohorts: all patients, 50-mile radius (DC/MD/VA), and DC residents. Predictors included age (defined as age at most recent encounter), sex, race/ethnicity, visit counts at each hospital, and year of most recent encounter. Continuous variables were standardized to per–standard deviation units. Year was included to account for secular trends.

Geographic disparities were examined at four scales: ward-level correlations (n = 8 wards, Pearson and Spearman), census tract–level correlations (n = 201 DC tracts and n = 698 regional tracts with ≥30 patients), zip code–level correlations (n = 130 zip codes), and individual-level neighborhood regression among DC residents. For individual-level regression, tract-level ACS data were linked directly to each patient’s geocoded census tract. Because the three neighborhood indicators measure overlapping dimensions of socioeconomic resources, they were entered into separate models rather than combined. As a sensitivity analysis, elastic net regularization (L1 + L2 penalty, 10-fold cross-validation) entered all three simultaneously to identify which retained independent predictive value; variance inflation factors for the neighborhood indicators ranged from 34 to 177 when combined, confirming severe multicollinearity. Variance inflation factors for individual-level predictors were all below 2.0. Within-ward racial disparities were examined by stratifying activation by race/ethnicity; subgroups with fewer than 30 patients were suppressed.

### Geospatial Visualization

Choropleth maps were generated using sf and ggplot2 in R. Regional maps display census tract–level data within an 8-mile square around central Washington, DC. Ward-level bar charts show activation and ACS indicators with scatterplots overlaid for visual correlation assessment. All analyses were conducted in R version 4.3 (R Foundation for Statistical Computing) with census tract geometries obtained via the tigris package.^20^

### Standard Protocol Approvals, Registrations, and Patient Consents

This study was reviewed and approved by the MedStar Health Research Institute Institutional Review Board (STUDY00007139) on October 19, 2023. The study was determined to be exempt (Category 4[iii]: secondary research on data or specimens) and the requirement for informed consent was waived. A waiver of HIPAA authorization was also granted.

### Data Availability

Individual patient data cannot be shared due to HIPAA restrictions on protected health information. Aggregate ward-level and census tract–level data are publicly available through the US Census Bureau American Community Survey. Anonymized aggregate data not published within this article may be made available by request from the corresponding author for purposes of replicating procedures and results.

## Results

### Study Population

Table 1 presents the study population across three nested geographic scales. Among all 72,417 patients, the mean age was 56.0 years (SD 18.8), 62.4% were female, and 38.9% were Non-Hispanic White. Most patients were seen at Hospital A and affiliated sites only (76.4%), with 20.1% at Hospital B only and 3.5% at both sites. Overall portal activation was 64.7%. Activation was higher among patients seen at both sites (87.2%) than Hospital A only (65.1%) or Hospital B only (59.5%).

**Table 1.** Study Population Characteristics Across Three Geographic Scales.

| Characteristic | All Patients<br>(N = 72,417) | 50-Mile Radius<br>(N = 69,071) | DC Residents<br>(N = 22,752) |
| --- | --- | --- | --- |
| **Portal Activated, n (%)** | 46,851 (64.7%) | 44,568 (64.5%) | 14,503 (63.7%) |
| Age, mean (SD) | 56.0 (18.8) | 56.2 (18.8) | 53.4 (18.6) |
| Female, n (%) | 45,182 (62.4%) | 43,119 (62.4%) | 14,772 (64.9%) |
| **Race/Ethnicity** |  |  |  |
| Non-Hispanic White | 28,154 (38.9%) | 26,299 (38.1%) | 6,594 (29.0%) |
| Non-Hispanic Black | 22,900 (31.6%) | 22,540 (32.6%) | 10,744 (47.2%) |
| Hispanic | 3,600 (5.0%) | 3,485 (5.0%) | 1,675 (7.4%) |
| Non-Hispanic Asian | 1,925 (2.7%) | 1,858 (2.7%) | 273 (1.2%) |
| Other/Unknown | 15,838 (21.9%) | 14,889 (21.6%) | 3,466 (15.2%) |
| **Hospital Site** |  |  |  |
| Hospital A only | 55,330 (76.4%) | 52,400 (75.9%) | 11,807 (51.9%) |
| Hospital B only | 14,525 (20.1%) | 14,193 (20.5%) | 9,542 (41.9%) |
| Both sites | 2,562 (3.5%) | 2,478 (3.6%) | 1,403 (6.2%) |
| **Jurisdiction** |  |  |  |
| DC | 22,752 (31.4%) | 22,752 (32.9%) | — |
| Maryland | 28,303 (39.1%) | 28,303 (41.0%) | — |
| Virginia | 18,016 (24.9%) | 18,016 (26.1%) | — |
| Other | 3,346 (4.6%) | — | — |

*50-mile radius includes DC, Maryland, and Virginia residents.*

### Activation Rates by Subgroup

Activation rates varied markedly by race/ethnicity across all scales (Table 2). Non-Hispanic White patients activated at 76.1% overall, compared to 57.0% for Non-Hispanic Black, 57.6% for Non-Hispanic Asian, and 55.0% for Hispanic patients. These disparities amplified within DC: Non-Hispanic White 87.1%, Non-Hispanic Black 51.0%, Hispanic 44.0%, while Non-Hispanic Asian patients in DC activated at 82.1%—though this subgroup was small (n = 273). Hospital A achieved higher activation than Hospital B in all cohorts (67.3% vs. 52.6% overall), reflecting differences in patient demographics rather than portal infrastructure.

**Table 2.** Portal Activation Rates by Patient Subgroup Across Three Geographic Scales.

| Subgroup | All Patients | 50-Mile Radius | DC Residents |
| --- | --- | --- | --- |
| **Overall** | 64.7% (46,851/72,417) | 64.5% (44,568/69,071) | 63.7% (14,503/22,752) |
| **Race/Ethnicity** |  |  |  |
| Non-Hispanic White | 76.1% (21,420/28,154) | 76.1% (20,015/26,299) | 87.1% (5,744/6,594) |
| Non-Hispanic Black | 57.0% (13,057/22,900) | 56.9% (12,835/22,540) | 51.1% (5,488/10,744) |
| Hispanic | 55.0% (1,979/3,600) | 54.2% (1,888/3,485) | 44.0% (737/1,675) |
| Non-Hispanic Asian | 57.6% (1,109/1,925) | 57.1% (1,061/1,858) | 82.1% (224/273) |
| Other/Unknown | 58.6% (9,286/15,838) | 58.9% (8,769/14,889) | 66.6% (2,310/3,466) |
| **Hospital Site** |  |  |  |
| Hospital A only | 67.3% (37,248/55,330) | 67.4% (35,295/52,400) | 76.5% (9,034/11,807) |
| Hospital B only | 52.6% (7,647/14,525) | 52.1% (7,393/14,193) | 47.0% (4,485/9,542) |
| Both sites | 76.3% (1,956/2,562) | 75.9% (1,880/2,478) | 70.1% (984/1,403) |

### Individual-Level Predictors of Portal Activation

Multivariable logistic regression revealed consistent predictors across all three cohorts (Table 3). Age was the strongest predictor: older patients activated at substantially lower rates, with each SD increase in age associated with 39–40% lower odds of activation. Racial/ethnic disparities amplified from the full cohort to DC: Non-Hispanic Black aOR 0.46 overall vs. 0.19 in DC; Hispanic aOR 0.34 overall vs. 0.11 in DC; Non-Hispanic Asian aOR 0.47 overall vs. 0.66 in DC. Hospital A visit frequency was strongly associated with activation across all scales. Year was independently significant, confirming secular improvement.

**Table 3.** Multivariable Logistic Regression: Predictors of Portal Activation Across Three Geographic Scales.

| Predictor | All Patients aOR (95% CI) | 50-Mile Radius aOR (95% CI) | DC Residents aOR (95% CI) |
| --- | --- | --- | --- |
| Age (per SD) | 0.60 (0.59–0.61) | 0.60 (0.59–0.62) | 0.63 (0.61–0.65) |
| Male (ref: Female) | 0.70 (0.67–0.72) | 0.70 (0.68–0.73) | 0.65 (0.61–0.69) |
| NHB (ref: NHW) | 0.46 (0.44–0.48) | 0.46 (0.44–0.48) | 0.19 (0.17–0.20) |
| Hispanic (ref: NHW) | 0.34 (0.31–0.37) | 0.33 (0.30–0.35) | 0.11 (0.10–0.13) |
| NH Asian (ref: NHW) | 0.47 (0.42–0.52) | 0.46 (0.41–0.51) | 0.66 (0.47–0.91) |
| Other/Unk (ref: NHW) | 0.42 (0.40–0.44) | 0.42 (0.40–0.44) | 0.28 (0.25–0.31) |
| Hosp A visits (per SD) | 3.24 (3.08–3.41) | 3.15 (2.99–3.31) | 3.58 (3.16–4.04) |
| Hosp B visits (per SD) | 1.14 (1.12–1.16) | 1.13 (1.10–1.15) | 1.03 (1.00–1.05) |
| Year (per SD) | 1.17 (1.15–1.19) | 1.18 (1.16–1.20) | 1.21 (1.17–1.25) |

*All models adjusted for listed variables simultaneously. Continuous variables standardized (per SD). NHW = Non-Hispanic White; NHB = Non-Hispanic Black. All $p < 0.001$ except Hospital B in DC ( $p = 0.049$ ).*

### Geographic Variation by DC Ward

Among 22,536 DC residents assigned to wards, activation rates ranged from 48.0% in Ward 7 to 82.0% in Ward 2—a 34.0 percentage point gap (Supplementary Table S5; Figure 2A; Supplementary Table S2). Wards in Northwest DC (Wards 2, 3, 6) had rates of 74.3–82.0%, while wards east of the Anacostia River (Wards 7, 8) had rates below 51%. This geographic pattern is visually striking across the regional maps: portal activation, median income, educational attainment, and broadband access all decline along a west-to-east gradient across the metropolitan area, with the sharpest transitions occurring at the Anacostia River (Figure 1).

**Figure 1.**
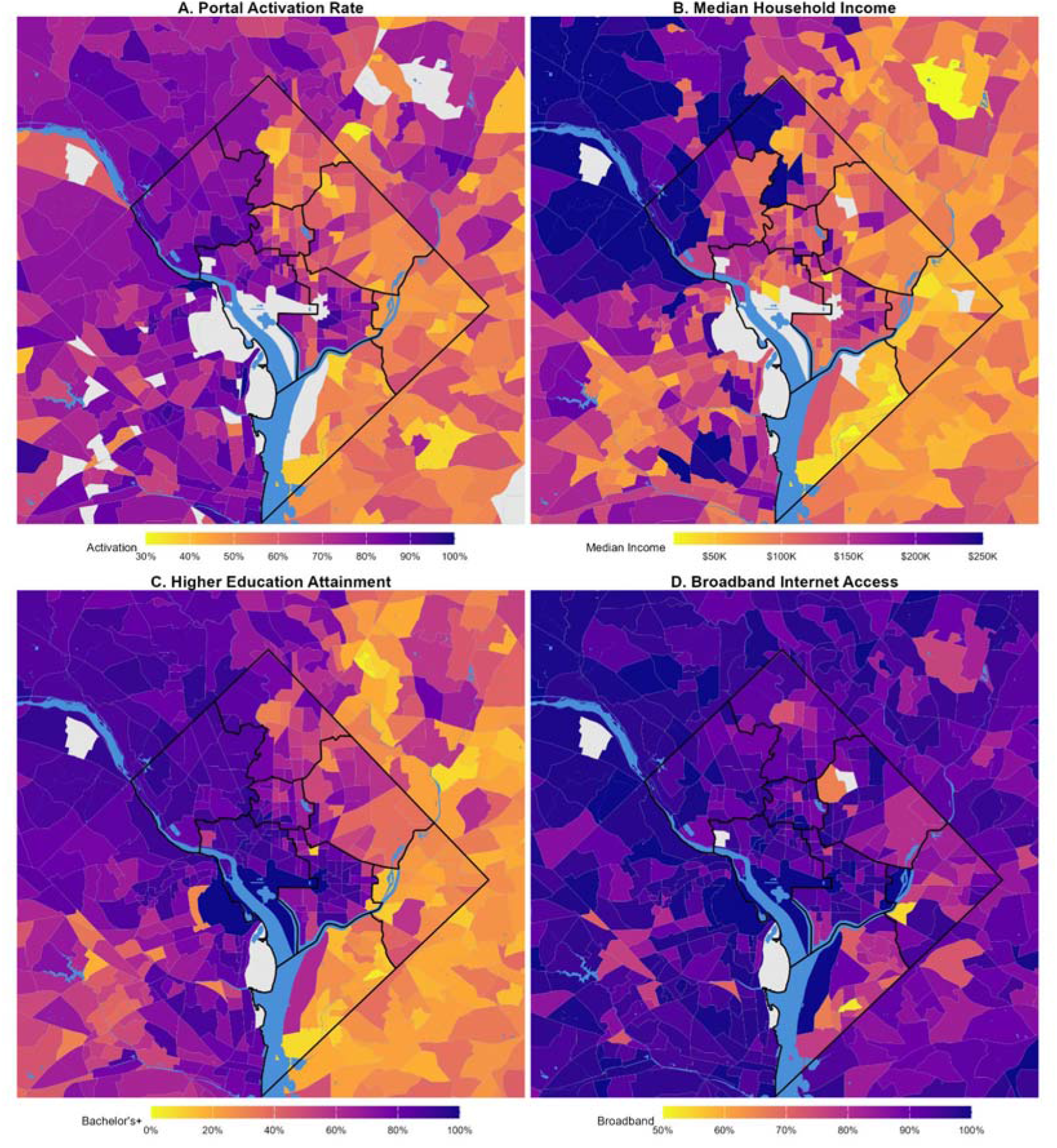
Regional Geographic Distribution of Portal Activation and Neighborhood Characteristics. 8-mile square around central DC. All panels: census tract level. Panel A: patient portal activation rate (tracts with <10 patients shown as gray). Panels B–D: ACS 2019–2023 5-year estimates. DC ward boundaries overlaid in black. Blue areas = water bodies. Gray areas = no data.

**Figure 2.**
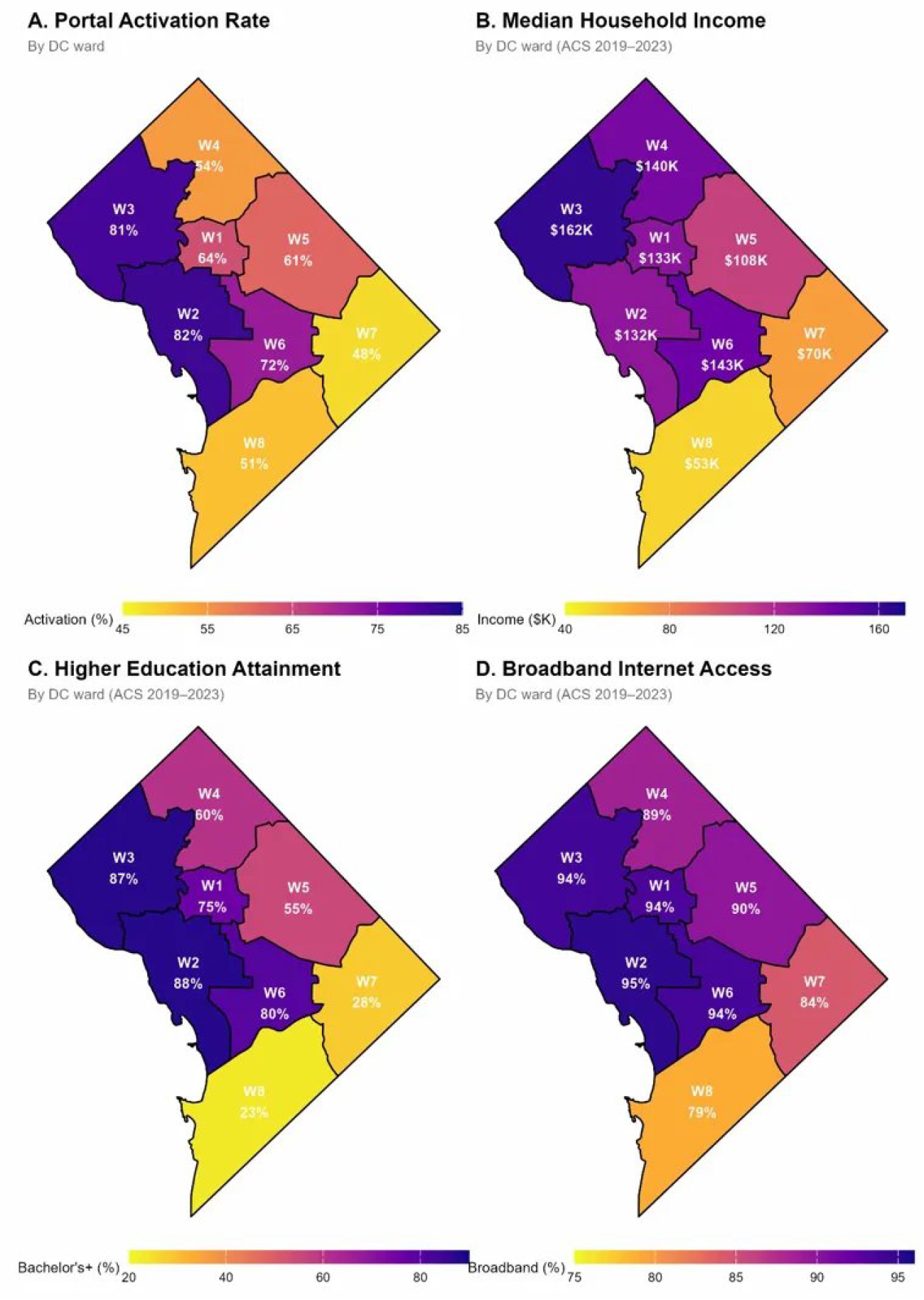
DC Ward-Level Portal Activation and Neighborhood Characteristics. N = 22,536 DC patients assigned via zip code + street address quadrant. (A) Portal activation by ward. (B) Median household income by ward. (C) Higher education attainment by ward. (D) Broadband internet access by ward. ACS 2019–2023 5-year estimates for DC wards (state legislative districts, upper chamber).

### Ecological Correlations

Ward-level correlations were strong and significant (Figure 2B–D): educational attainment (r = 0.948, p < 0.001), broadband access (r = 0.889, p = 0.003), and median income (r = 0.811, p = 0.015). Census tract–level correlations among 201 DC tracts with ≥30 patients confirmed these patterns: education (r = 0.853, p < 0.001), income (r = 0.658, p < 0.001), and broadband (r = 0.598, p < 0.001). Among 698 tracts across the full regional catchment, correlations were attenuated but remained significant: education (r = 0.418), income (r = 0.252), broadband (r = 0.221; all p < 0.001). Zip-level correlations were weakest (Supplementary Table S1). Spearman correlations confirmed these patterns at all scales.

### Within-Ward Racial Disparities

Racial disparities persisted within individual wards. In Ward 6 (income $ 143,000; 80% bachelor’s), Non-Hispanic White patients activated at 90.9% vs. 53.4% for Non-Hispanic Black—a 37.5-point gap. Across Wards 1–6, Non-Hispanic White activation ranged from 83.7% to 90.9% while Non-Hispanic Black ranged from 47.9% to 63.6%. This demonstrates that neighborhood resources alone do not explain disparities.

### Neighborhood-Level Predictors

Among DC residents (N = 22,536), each ward-level socioeconomic variable was significantly associated with activation after demographic adjustment (Supplementary Table S3). Income: aOR 1.18 (1.14–1.21); education: aOR 1.26 (1.22–1.30); broadband: aOR 1.22 (1.18– 1.26) per SD. When tract-level ACS data were linked directly to individual patients via geocoded addresses (N = 21,567), associations were consistently stronger: income aOR 1.31 (1.26–1.35); education aOR 1.38 (1.33–1.43); broadband aOR 1.22 (1.18–1.26) per SD. Substantial attenuation from unadjusted to adjusted estimates (e.g., tract income from OR 1.59 to 1.31) reflects overlap between neighborhood characteristics and patient race/ethnicity. Elastic net regularization with all three indicators entered simultaneously confirmed that broadband access was subsumed by income and education.

### Temporal Trends

Patients whose most recent encounter was in 2026 had higher activation (79.3%) than those last seen in 2021 (55.4%), consistent with both secular improvement in portal adoption and selective retention of engaged patients. This temporal gradient was evident across racial groups but did not close disparities: Non-Hispanic White patients improved from 64.2% (2021) to 87.1% (2026) while Non-Hispanic Black patients improved from 48.3% to 72.1%, maintaining a persistent 15-point gap. Year as a continuous predictor was significant in all regression models (aOR 1.17, 95% CI 1.15–1.19; Supplementary Table S4).

## Discussion

This analysis of 72,417 neurology patients shows that portal activation is shaped by converging demographic, socioeconomic, and geographic factors at both individual and neighborhood levels. The three-cohort design reveals how disparities emerge at one geographic scale and deepen at the next.

Older age was the strongest individual predictor of non-activation across all cohorts (aOR 0.60–0.63 per SD), consistent with national trends in digital health adoption.^11,13^ Racial disparities amplified as the geographic lens narrowed: the Non-Hispanic Black aOR was 0.46 overall but 0.19 among DC residents; the Hispanic aOR was 0.34 overall but 0.11 in DC. Non-Hispanic Asian patients showed a different pattern (aOR 0.47 overall, 0.66 in DC), suggesting their activation disadvantage is attenuated within the District. These deepening odds ratios likely reflect the residential concentration of minority patients in under-resourced eastern wards.^21,22^

The 34.0-point gap between Ward 2 (82.0%) and Ward 7 (48.0%) mirrors DC’s well-documented health inequities, where life expectancy differs by roughly 15 years across wards.^21^ All three socioeconomic indicators correlated strongly with activation (r = 0.81–0.95), with educational attainment showing the strongest association at both ward and tract levels— suggesting it captures dimensions of digital readiness that income alone does not.

Perhaps the most consequential finding is that neighborhood advantage does not resolve individual disadvantage. Racial gaps of 30–40 percentage points persisted within individual wards, even the most affluent. In Ward 6—where median income exceeds $ 143,000 and 80% of adults hold college degrees—Non-Hispanic White patients activated at 90.9% compared to 53.4% for Non-Hispanic Black patients. This pattern held across all six wards where comparison was possible.

These within-ward findings represent, to our knowledge, the first demonstration that racial disparities in portal activation persist independent of neighborhood socioeconomic advantage. Prior studies documented racial gaps at the population level using national survey d^5,24^5,24 but lacked the geographic resolution to test whether neighborhood resources mitigate individual-level disparities. By geocoding 94.8% of DC patients to census tracts and linking them to tract-level ACS data, this study localizes where within communities these gaps concentrate.

The neighborhood regression supports this: after demographic adjustment, tract-level income attenuated from OR 1.59 to 1.31, indicating that much of the apparent neighborhood effect reflects its correlation with patient race and ethnicity. Tract-level ACS linkage yielded stronger associations than ward-level averages (e.g., education aOR 1.38 vs. 1.27), consistent with finer socioeconomic variation at the tract level. Neighborhood disadvantage and individual disparities are not independent forces but mutually reinforcing ones.^9,10^

The multi-scale correlation analysis illustrates how geographic aggregation and population composition shape observed associations. Ward-level correlations were strongest (r = 0.81–0.95) partly because averaging across ~25 census tracts per ward smooths within-ward variation. Among 201 DC tracts, finer resolution revealed heterogeneity masked by ward averages (r = 0.60–0.85). Among 698 regional tracts, correlations attenuated further (r = 0.22– 0.42), reflecting both disaggregation and the inclusion of demographically distinct suburban populations. Zip-level correlations were weakest (r = 0.16–0.39). Together, these results show that the socioeconomic–activation relationship is robust, but its apparent strength depends on the level of aggregation and the composition of the analytic sample.

This multi-scale approach is novel in the portal disparities literature, where prior studies have typically operated at a single geographic level.24 For neurological patients managing complex, chronic conditions, portal access supports medication management, appointment coordination, and test result review—functions for which access remains unequally distributed.6 The strong broadband–activation correlation (r = 0.889) underscores this point, as mobile portal access has been independently linked to improved medication adherence.7,14

Portal activation is not an unqualified good. Portals can fragment the patient–provider relationship when asynchronous messaging replaces direct conversation, and overreliance on portal communication may disadvantage patients with complex neurological conditions. Higher activation also increases clinician inbox volume—largely uncompensated under current reimbursement models—contributing to burnout. The goal is equitable access to a tool that, when integrated thoughtfully, extends rather than replaces the therapeutic relationship.

## Limitations

Portal activation was binary (account status) rather than measuring actual use. The retrospective design precludes causal inference. Year of most recent encounter conflates temporal trends with patient engagement; patients last seen in earlier years may include those who transferred care or were lost to follow-up, and their lower activation rates may partly reflect disengagement from the health system rather than historical adoption patterns.

Ward assignment via zip code and quadrant remains approximate near boundaries, though census tract geocoding of 94.8% of DC addresses provides complementary precision; the 6.1% not geocoded were demographically similar to geocoded patients, suggesting minimal selection bias. The ecological design is subject to ecological fallacy; however, the 201-tract and 130-zip analyses demonstrate that the socioeconomic–activation relationship holds at finer geographic resolution.

The three ACS neighborhood indicators were intercorrelated (r > 0.89 at ward level), as expected given overlapping socioeconomic dimensions; accordingly, they were examined in separate models. Patient data (2021–2026) and ACS estimates (2019–2023) are partially temporally mismatched, though persistent structural disparities make this unlikely to affect findings meaningfully. Broadband subscription rates do not capture internet quality, digital literacy, or device access. The Other/Unknown race/ethnicity category (21.9%) may obscure additional disparities.

## Conclusions

Disparities in patient portal activation among neurology patients are multifactorial. Age, race, neighborhood context, and healthcare system factors each contribute independently but compound in combination. A younger, Non-Hispanic White patient in Ward 2 activates at 91%; an older, Non-Hispanic Black patient in Ward 7, at 36%. That 55-point gap is not the product of any single factor but the accumulation of all of them.

No single intervention will close this gap. Infrastructure investment alone will not overcome barriers rooted in digital literacy. Culturally tailored outreach alone will not compensate for neighborhoods lacking broadband. Closing the digital divide requires coordinated approaches—targeted to the populations and geographies where the divide is widest—addressing structural, demographic, and clinical barriers simultaneously.

This analysis maps where the digital divide in healthcare runs deepest. The geographic and demographic granularity presented here provides a foundation for intervention design. A natural next step is individual-level prediction—integrating these variables into models that flag patients at highest risk of non-activation, enabling proactive outreach before patients fall through the digital divide.

## Acknowledgments

The authors wish to thank Mindi Messmer, DMSc, PG, CG (Senior Research Scientist, MedStar Health Research Institute, Center for Biostatistics, Informatics, and Data Science [CBIDS]), for her contributions to statistical analysis and data management, and Jiling Chou (MedStar Health Research Institute, CBIDS) for analytical support.

