## Supplemental Tables for "Patient Portal Activation Among Neurology Patients in Washington, DC" (from another version of this preprint: version 1, 2026-04-09)

**Supplementary Materials**

*Geographic and Demographic Determinants of Patient Portal Access: A Multi-Level Analysis of 72,417 Neurology Patients*

**Supplementary Table S1. Multi-Scale Ecological Correlations with Pearson and Spearman Coefficients**

Ecological correlations between neighborhood socioeconomic characteristics and portal activation rates at four geographic scales. Spearman correlations are provided for the ward level to account for the small sample size (n = 8) and potential outlier influence.

| **Scale** | **N** | **Income** | **Education** | **Broadband** |
| --- | --- | --- | --- | --- |
| **DC Wards** |  |  |  |  |
| Pearson r | 8 | 0.811 (p=.015) | 0.948 (p<.001) | 0.889 (p=.003) |
| Spearman ρ | 8 | 0.833 (p=.010) | 0.929 (p=.001) | 0.952 (p<.001) |
| **DC Tracts (≥30)** |  |  |  |  |
| Pearson r | 201 | 0.658 (p<.001) | 0.853 (p<.001) | 0.598 (p<.001) |
| **Regional Tracts (≥30)** |  |  |  |  |
| Pearson r | 698 | 0.252 (p<.001) | 0.418 (p<.001) | 0.221 (p<.001) |
| **Zip/County (≥30)** |  |  |  |  |
| Pearson r | 130 | 0.221 (p=.012) | 0.385 (p<.001) | 0.160 (p=.068) |

*Pearson r and Spearman ρ with exact p-values. Ward: activation vs. ACS 2019–2023 ward-level estimates (state legislative districts, upper chamber). Tract: activation vs. tract-level ACS among tracts with ≥30 patients. Zip: activation vs. county-aggregated ACS for zip codes with ≥30 patients.*

**Supplementary Table S2. DC Ward Characteristics: Census Tract Composition and ACS Estimates**

ACS 2019–2023 5-year estimates obtained directly at the ward level via tidycensus (DC wards = state legislative districts, upper chamber, in Census geography).

| **Ward** | **Tracts** | **Income** | **Bachelor’s+** | **Broadband** | **N Patients** | **Activation** |
| --- | --- | --- | --- | --- | --- | --- |
| 1 | 22 | $126,000 | 76% | 93% | 2,985 | 68.9% |
| 2 | 28 | $131,000 | 87% | 95% | 1,896 | 82.0% |
| 3 | 23 | $142,000 | 87% | 93% | 2,831 | 80.7% |
| 4 | 23 | $128,000 | 60% | 89% | 4,264 | 54.9% |
| 5 | 26 | $98,000 | 55% | 90% | 3,874 | 61.1% |
| 6 | 35 | $138,000 | 82% | 94% | 1,895 | 74.3% |
| 7 | 25 | $69,000 | 32% | 85% | 2,131 | 48.0% |
| 8 | 24 | $51,000 | 29% | 81% | 2,660 | 50.8% |
| **Total** | **206** | — | — | — | **22,536** | **63.8%** |

*ACS values obtained directly at the ward level from the Census Bureau. Tract counts reflect number of DC census tracts per ward. Income = median household income. Bachelor’s+ = percentage of adults ≥25 with bachelor’s degree or higher. Broadband = percentage of households with broadband subscription.*

**Supplementary Table S3. Individual-Level Neighborhood Regression: Ward vs. Tract ACS Linkage (Full Model Detail)**

Adjusted odds ratios from separate logistic regression models, each including one neighborhood socioeconomic variable plus patient age, sex, and race/ethnicity. Neighborhood variables standardized to per–standard deviation units. Ward-level models: N = 22,536 DC residents. Tract-level models: N = 21,335 geocoded DC residents.

| **Predictor** | **Income Model aOR (95% CI)** | **Education Model aOR (95% CI)** | **Broadband Model aOR (95% CI)** |
| --- | --- | --- | --- |
| **Neighborhood (per SD)** |  |  |  |
| Ward-level ACS | 1.18 (1.14–1.21) | 1.26 (1.22–1.30) | 1.22 (1.18–1.26) |
| Tract-level ACS | 1.31 (1.26–1.35) | 1.38 (1.33–1.43) | 1.22 (1.18–1.26) |
| **Patient Covariates** |  |  |  |
| Age (per SD) | 0.65 (0.63–0.67) | 0.65 (0.63–0.67) | 0.65 (0.63–0.67) |
| Male (ref: Female) | 0.63 (0.59–0.67) | 0.63 (0.59–0.67) | 0.63 (0.59–0.67) |
| NHB (ref: NHW) | 0.17 (0.16–0.19) | 0.19 (0.18–0.21) | 0.18 (0.17–0.20) |
| Hispanic (ref: NHW) | 0.10 (0.08–0.11) | 0.10 (0.09–0.12) | 0.10 (0.09–0.12) |
| NH Asian (ref: NHW) | 0.59 (0.43–0.82) | 0.61 (0.44–0.84) | 0.60 (0.43–0.83) |
| Other/Unk (ref: NHW) | 0.26 (0.23–0.29) | 0.27 (0.24–0.30) | 0.27 (0.24–0.30) |

*Each column represents a separate model with the specified neighborhood variable. All neighborhood and patient variables are mutually adjusted within each model. Ward ACS = ACS 2019–2023 ward-level estimates (state legislative districts). Tract ACS = ACS 2019–2023 linked directly to patient’s geocoded census tract. All p < 0.001.*

**Supplementary Table S4. Portal Activation Rates by Year and Race/Ethnicity Across Three Geographic Scales**

Activation rates stratified by year of most recent encounter and race/ethnicity. The NHW–NHB Gap column shows the percentage point difference between Non-Hispanic White and Non-Hispanic Black activation rates within each year.

**Panel A: All Patients**

| **Year** | **Overall** | **NHW** | **NHB** | **Hispanic** | **NH Asian** | **Other/ Unk** | **NHW–NHB Gap** |
| --- | --- | --- | --- | --- | --- | --- | --- |
| 2021 | 55.4% | 67.3% | 47.1% | 42.2% | 50.0% | 53.3% | 20.2 |
| 2022 | 59.5% | 72.2% | 51.8% | 48.3% | 62.6% | 52.6% | 20.3 |
| 2023 | 57.6% | 67.8% | 52.2% | 55.3% | 44.6% | 49.9% | 15.6 |
| 2024 | 61.1% | 71.5% | 55.6% | 52.7% | 48.2% | 55.8% | 15.9 |
| 2025 | 70.0% | 81.0% | 63.6% | 59.2% | 66.5% | 62.4% | 17.4 |
| 2026* | 79.3% | 89.2% | 68.8% | 69.9% | 81.5% | 72.0% | 20.4 |
| **Total** | **64.7%** | **76.1%** | **57.0%** | **55.0%** | **57.6%** | **58.6%** | **19.1** |

**2026 includes January–February only. NHW = Non-Hispanic White; NHB = Non-Hispanic Black.*

**Panel B: 50-Mile Radius (DC/MD/VA)**

| **Year** | **Overall** | **NHW** | **NHB** | **Hispanic** | **NH Asian** | **Other/ Unk** | **NHW–NHB Gap** |
| --- | --- | --- | --- | --- | --- | --- | --- |
| 2021 | 54.5% | 66.3% | 46.9% | 41.1% | 49.2% | 53.1% | 19.4 |
| 2022 | 58.8% | 71.6% | 51.4% | 47.4% | 61.5% | 52.6% | 20.1 |
| 2023 | 57.4% | 68.1% | 52.4% | 54.0% | 43.3% | 49.9% | 15.7 |
| 2024 | 61.0% | 71.6% | 55.6% | 52.1% | 48.1% | 55.8% | 16.0 |
| 2025 | 69.9% | 80.9% | 63.5% | 58.4% | 66.2% | 62.9% | 17.5 |
| 2026* | 79.2% | 89.1% | 68.8% | 69.8% | 81.5% | 71.9% | 20.4 |
| **Total** | **64.5%** | **76.1%** | **56.9%** | **54.2%** | **57.1%** | **58.9%** | **19.2** |

**2026 includes January–February only.*

**Panel C: DC Residents**

| **Year** | **Overall** | **NHW** | **NHB** | **Hispanic** | **NH Asian** | **Other/ Unk** | **NHW–NHB Gap** |
| --- | --- | --- | --- | --- | --- | --- | --- |
| 2021 | 52.7% | 79.3% | 41.2% | 29.3% | 81.2% | 57.0% | 38.2 |
| 2022 | 57.1% | 83.4% | 45.4% | 34.2% | 77.8% | 58.8% | 38.0 |
| 2023 | 60.0% | 83.2% | 48.5% | 44.6% | 65.9% | 61.9% | 34.7 |
| 2024 | 61.5% | 85.8% | 51.1% | 39.1% | 83.6% | 62.0% | 34.7 |
| 2025 | 69.4% | 90.8% | 56.7% | 51.1% | 87.7% | 73.1% | 34.1 |
| 2026* | 77.1% | 93.6% | 60.2% | 67.2% | 96.3% | 78.1% | 33.4 |
| **Total** | **63.7%** | **87.1%** | **51.1%** | **44.0%** | **82.1%** | **66.6%** | **36.0** |

**2026 includes January–February only. The NHW–NHB gap among DC residents (38.1 to 33.4 points) is substantially larger than in the full cohort (20.2 to 20.4 points), reflecting the concentration of Non-Hispanic Black patients in under-resourced eastern wards. Although absolute activation rates improved for all groups, the persistent gap indicates that secular improvement alone is insufficient to achieve equity.*

**Supplementary Table S5. Portal Activation and Neighborhood Characteristics by DC Ward**

| **Ward** | **N** | **Activation** | **Income** | **Bachelor's+** | **BB** | **%NHB** | **NHW Act.** | **NHB Act.** | **Hispanic Act.** | **NH Asian Act.** |
| --- | --- | --- | --- | --- | --- | --- | --- | --- | --- | --- |
| 1 | 2,985 | 68.9% | $126K | 76% | 93% | 34% | 88.6% | 51.3% | 47.8% | 80.4% |
| 2 | 1,896 | 82.0% | $131K | 87% | 95% | 10% | 87.3% | 59.2% | 76.0% | 87.2% |
| 3 | 2,831 | 80.7% | $142K | 87% | 93% | 11% | 85.7% | 63.6% | 69.6% | 87.7% |
| 4 | 4,264 | 54.9% | $128K | 60% | 89% | 47% | 86.1% | 52.7% | 29.1% | 73.7% |
| 5 | 3,874 | 61.1% | $98K | 55% | 90% | 58% | 87.8% | 50.5% | 52.7% | 85.4% |
| 6 | 1,895 | 74.3% | $138K | 82% | 94% | 38% | 91.2% | 53.6% | 76.9% | 81.8% |
| 7 | 2,131 | 48.0% | $69K | 32% | 85% | 87% | 66.7% | 47.6% | 30.9% | * |
| 8 | 2,660 | 50.8% | $51K | 29% | 81% | 86% | 72.1% | 49.7% | 42.9% | * |
| **Total** | 22,536 | 63.8% | **—** | **—** | **—** | **—** | **—** | **—** | **—** | **—** |

**Subgroup n < 30, suppressed. ACS 2019–2023 census tracts spatially joined to wards. BB = broadband. NHW/NHB activation rates shown where n ≥ 30.*
